# Importance of serological testing in the convalescence phase in patients with pulmonary impairment due to COVID 19 - a health care workers analysis

**DOI:** 10.1101/2021.03.24.20208835

**Authors:** José Rodrigues Pereira, Ilka Lopes Santoro, Maria Silvia Biagioni Santos, Andreia Padilha de Toledo, Greice Elen Copelli, Caroline Villela Galvão de França, Edmundo Di Giaimo Caboclo

## Abstract

Since its discovery, more than 37 million people have been infected by SARS-CoV-2 with deaths around 1 million worldwide. The prevalence is not known because infected individuals may be asymptomatic. In addition, the use of specific diagnostic tests is not always conclusive, raising doubts about the etiology of the disease.

The best diagnostic method and the ideal time of collection remains the subject of study. The gold standard for diagnosing COVID 19 is the RT PCR molecular test, usually using an oropharynx and nasopharynx swab. Its sensitivity is 70% and drops significantly after the second week of symptoms. Serological tests, in turn, have increased sensitivity after 14 days, and can contribute to the diagnosis when SARS-CoV-2 infection is suspected, even with negative RT PCR.

Our study showed sensitivity and specificity of 100% of the serological test (ELISA method) for cases of viral pneumonia caused by the new coronavirus, suggesting that this test could assist in the diagnosis of pulmonary interstitial changes that have not yet been etiologically clarified. We found a greater immune response in men, regardless of the severity of symptoms. The greater the severity, the higher the levels of IgA and IgG, mainly found in patients with multilobar impairment and in need for oxygen. We concluded that the serological test collected around 30 days after the onset of symptoms is the best diagnostic tool in the convalescence phase, not only for epidemiological purposes, but also for the etiological clarification of pulmonary changes that have not yet been diagnosed.

## 1 Introduction

Since the very first case of SARS-CoV-2 in December 2019 until the October 13, 2020, the cumulative total of cases worldwide was 37,704,153 and 1,079,029 deaths. In Brazil, the number of cases and deaths by COVID 19 correspond to 5,094,979 and 150,488, respectively (1-2). According to the World Health Organization (WHO), cases of COVID 19 confirmed through RT-PCR and who have pulmonary impairment are classified, at least as moderate COVID. This classification can change depending on the clinical repercussion of pulmonary damage, in other words, the need for supplemental oxygen supply due to hypoxemia or respiratory rate ≥ 30 bpm; the need for mechanical ventilation, lead to COVID 19 classification as severe or critical, respectively (3).

The ideal moment for carrying out diagnostic tests has been questioned, and the most specific test is the RT-PCR. Despite being the gold standard for diagnosis, its sensitivity varies according to the time of collection concerning the onset of symptoms, appropriate technique, and representativeness of the material, which can result in a low percentage of positivity (4). Serological tests for COVID 19 could help in the diagnosis, but due to the low sensitivity in the acute phase, its use is more accurate 14 days after the onset of symptoms, when the infected patient has already gone through the viral cycle. Evidence in the literature has shown an increase in the sensitivity of the serological test when the severity of the disease is greater (5).

Another exam used in the clinical suspicion of COVID 19 is Chest Computerized Tomography (CT), which has good sensitivity in assessing pulmonary disease extend possible differential diagnoses or complications (6). The main tomographic finding in the initial phase of the disease is bilateral ground-glass opacities, predominantly peripheral and in the lower fields (7-10). As the disease progresses, from the 7th day onwards, opacities acquire a consolidating characteristic (11).

The combination of tomographic findings with serological tests could enhance the sensitivity of the two exams, improving diagnostic accuracy.

This study aims to evaluate the sensitivity and specificity of serology in patients with the pulmonary manifestation of COVID 19 after the end of the infectious cycle, to establish another pattern of diagnosis in the phase of convalescence in which there were a pulmonary repercussion and persistence of sequels. As a secondary objective, to assess differences in the titration of specific acute-phase (IgA) and late-phase (IgG) antibodies according to the time of symptom onset, sex, severity, comorbidities, and persistence of positive RT PCR.

### 2 Materials and methods

Data were analyzed from 26 immunocompetent employees of Hospital Beneficência Portuguesa de São Paulo, who had a diagnosis of COVID 19 confirmed through RT-PCR (molecular analysis collected through an oropharynx and nasopharynx swab) or suspected due to the presence of pneumonia defined by the clinical condition and change in chest CT. All evaluated patients required hospitalization, from April 1st to May 31st, 2020, and after hospital discharge, they underwent serological tests.

Retrospective analysis of the medical records was performed at the time of hospitalization and / or outpatient care.

The following clinical variables were evaluated: age (stratified 10-year age groups from 20 to 60 years old or more), sector of work in the hospital, sex, first symptom, pneumonia symptoms (cough, dyspnea, and fever), comorbidities (classified according to presence and type), length of hospital stay, positive 1st and 2nd RT-PCR for COVID 19, positive serology for COVID 19 and extent of pneumonia (multilobar or unilobar). A list of antibody levels (IgA and IgG) was also performed according to sex, collection time from symptom onset (≤ 4 weeks or> 4 weeks), presence of the need for supplemental oxygen, multilobar impairment, comorbidities, the severity of lung disease, and positivity of the 2nd RT PCR.

The serological method used was ELISA (Enzyme-Linked ImmunoSorbent Assay), with results <0.8 - negative; 0.8 to 1.09 - intermediate and> 1.10 - positive, both for Immunoglobulin A (IgA) and for Immunoglobulin G (IgG).

### 3.1 Statistical analysis

For statistical analysis, categorical variables were expressed in absolute numbers and percentages, assessed by the chi-square test or Fisher’s exact test. Continuous data were expressed as mean (standard deviation) or median (interquartile range), depending on the nature of the variable. The groups were compared using the Mann-Whitney or Kruskal Wallis test, due to the nature of the dependent variable. Statistical analysis was performed using SPSS v19 software.

## 4 Results

In this sample, it was observed that the median age was 40.5 years with the majority of patients aged from 30 to 49 years, corresponding to 66% of the total number of individuals evaluated (17/26). There was a predominance of ICU employees (17/26) and a predominance of females (18/26). All patients suspected or confirmed for COVID 19 through positive RT PCR presented positive serological tests (26/26), with a collection median of 31.5 days from the onset of symptoms. The predominant comorbidities among those hospitalized by COVID-19 were obesity and arterial hypertension. Besides, hospitalization was mainly earlier (< 5 days) and patients were with multilobar lung injuries. (Table 1).

**Table 1.**
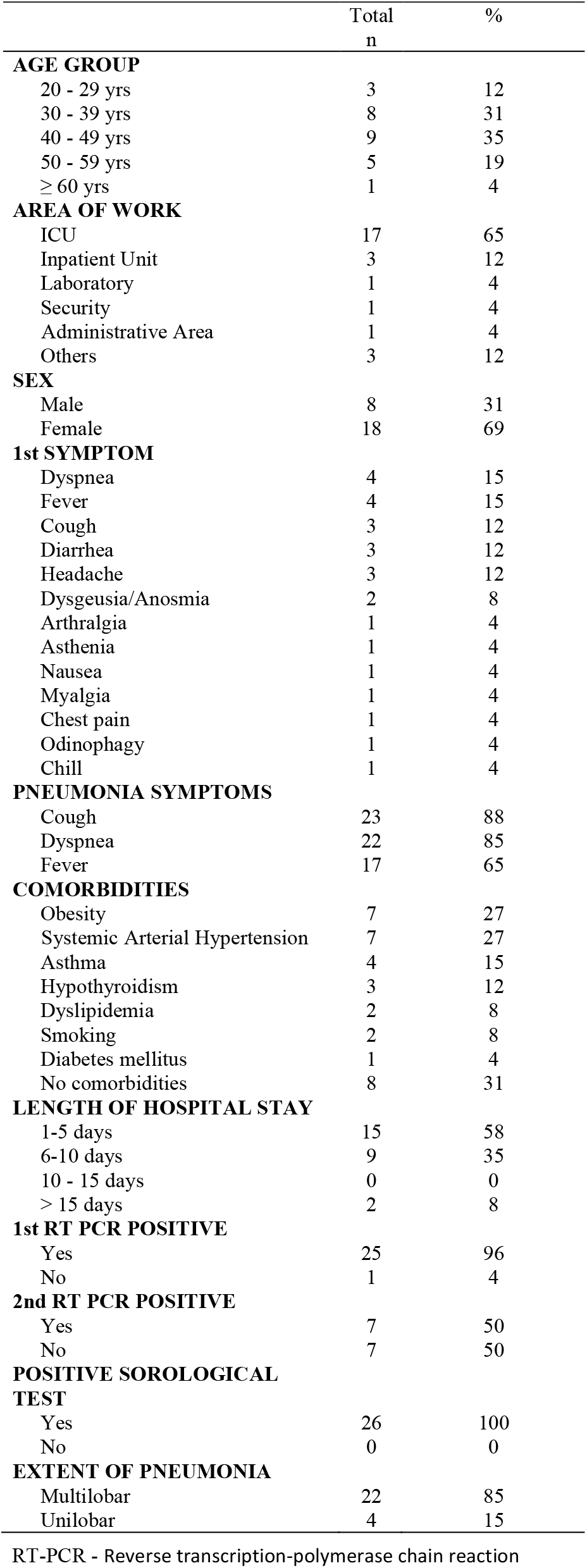
Characteristics of Patients

The levels of antibodies were, for IgA, higher in men (6x higher levels), as well as in multilobar impairment. We also observed a decrease in IgA values and an increase of IgG when collected from the day 28 (Table 2).

**Table 2.** Antibody Titration X Clinical Characteristics

|  | IgA md [IQR] | Total n 25 (%) | P Value | IgG md [IQR] | Total n 26 (%) | P Value |
| --- | --- | --- | --- | --- | --- | --- |
| **SEX** |  |  |  |  |  |  |
| Male | 34.2 [4.8 – 39.86] | 8 (32%) | **0.029** | 11.15 [6.7 – 16.87] | 8 (31%) | 0.107 |
| Female | 6.53 [0.77 – 34.20] | 17 (68%) |  | 8.17 [1.3 – 14.4] | 18 (69%) |  |
| **WEEKS** |  |  |  |  |  |  |
| ≤ 4 Weeks | 12.02 [2.42 – 34.2] | 9 (36%) | 0.977 | 6.42 [1.3 – 14.24] | 10 (38%) | **0.035** |
| > 4 Weeks | 6.9 [0.77 – 40.2] | 16 (64%) |  | 11.27 [4.54 -17.95] | 16 (62%) |  |
| **NEED FOR O2** |  |  |  |  |  |  |
| Yes | 33.47 [2.42 – 34.93] | 9 (36%) | 0.21 | 13.26 [4.09 – 14.63] | 9 (35%) | **0.019** |
| No | 6.85 [0.77 -39.26] | 16 (64%) |  | 7.99 [1.30 – 13.47] | 17 (65%) |  |
| **MULTILOBAR** |  |  |  |  |  |  |
| Yes | 12.54 [2.64 – 40.2] | 21 (84%) | **0.012** | 11.05 [1.68 - 18.46] | 22 (85%) | **0.023** |
| No | 2.56 [0.77 – 5.89] | 4 (16%) |  | 5.18 [2.44 – 6.51] | 4 (15%) |  |
| **COMORBIDITIES** |  |  |  |  |  |  |
| Yes | 12.65 [1.14 -38.94] | 18 (72%) | 0.74 | 11.05 [1.3 – 15.10] | 18 (69%) | 0.134 |
| No | 5.16 [0.77 – 5.73] | 7 (28%) |  | 6.42 [3.8 – 11.34] | 8 (31%) |  |

IgA- Immunoglobulin A; IgG - Immunoglobulin G; md - Median; IQR -Interquartile range

The greater severity of the condition, characterized by multilobar impairment and the need for supplemental oxygen supply, was associated with higher levels of IgA and IgG (Graph 1).

Out of the patients who showed persistence of the 2nd positive RT PCR, with a median collection time of 30 days, we did not observe a greater immune response. (Table 3)

**Table 3.** Antibody Titration x 2nd RT PCR

| 2nd RT PCR<br>Positive | IgA md [IQR] | Total n 13 (100%) | P Value | IgG md [IQR] | Total n 14 (100%) | P Value |
| --- | --- | --- | --- | --- | --- | --- |
| Yes | 5.73 [1.14 – 9.60] | 7 (54%) | 0.253 | 7.16 [3.86 – 11.05] | 7 (50%) | 0.482 |
| No | 9.69 [3.99 – 33.47] | 6 (46%) |  | 11.87 [2.44 – 12.13] | 7 (50%) |  |

RT-PCR - Reverse transcription-polymerase chain reaction; IgA - Immunoglobulin A; IgG - Immunoglobulin G; md - Median; IQR - Interquartile range;

## 5 Discussion

The gold standard for the diagnosis of COVID 19 is the molecular analysis of material from the upper or lower airways using RT-PCR. As an auxiliary diagnostic tool, specific serological tests with characterization and quantification of total antibodies, neutralizing antibodies, IgM and IgA (acute phase antibodies), and IgG (late phase antibody), have been extensively studied. Its real relevance and interpretation of results have still generated discussion in the scientific literature and studies comparing different methodologies have evaluated its sensitivity and specificity (12-13).

In our study, the serological test using the ELISA method was positive in 100% of patients with confirmed or suspected pulmonary impairment for COVID 19, when performed 14 days after the onset of symptoms (26/26).

A patient with symptoms, characteristic tomographic presentations, and persistently negative RT-PCR (two collections on different days), also presented positive serology, confirming the diagnosis of infection with the new coronavirus. Patients with suspected clinical and tomographic changes for COVID 19 may have repeatedly negative RT-PCR results (6). The sensitivity of the test decreases after the second week of symptom onset, allowing patients infected with SARS-CoV-2, by successive negative RT-PCRs, to be erroneously characterized as having another disease (14).

Evidence in the literature shows that chest CT would have a higher sensitivity for detecting SARS-CoV-2 pneumonia compared to molecular tests in the early stage of the disease (15). For sensitivity of RT PCR when performed between the first to the seventh day after the onset of symptoms is 88.9%, 73.7%, and 60% for sputum, nasopharynx, and oropharynx swab, respectively. In mild cases for the same time interval, the sensitivity is 82.2%, 72.1%, and 61.3%, respectively. When the time interval is from the eighth to the fourteenth day of symptom onset, in severe cases the positivity is 100% in bronchoalveolar lavage (BAL), 83.3% in sputum, 72.3% in the nasal swab, and 50% in the oropharynx swab. For mild cases, after the eighth day, detection is absent for BAL and 74.4%, 53.6% and 29% for sputum, nasal and oropharyngeal swab (16). Thus, the most practical and representative material would be the nasal swab for both mild and severe cases, but with the caveat that its sensitivity is around 70%. A systematic review of the literature showed that the rate of false-negative results from RT-PCR is 29% (6). Out of the patients evaluated, 96% were positive for IgA (24/25) and 100% were positive for IgG (26/26), both of which were collected after 14 days. K Imai et al., using the immunochromatography technique, found IgM and IgG sensitivity for symptomatic patients with more than 14 days of symptoms of 95.8% and 65.2%, respectively. When the serological test is combined with chest tomography in symptomatic patients, sensitivity after two weeks reaches 100% (17).

As the molecular analysis through RT-PCR can be falsely negative, the erroneous investigative conduct of the case would lead to misdiagnosis with new exams and unnecessary treatments, thus postponing fundamental interventions to prevent the patient from evolving with pulmonary fibrosis and permanent functional alteration.

Patients confirmed for COVID 19 who presented pneumonia maintained presented pneumonia maintained evaluated around 30 days after the onset of symptoms, reaching 84.2% of DLCO in the pulmonary function test. (18)

**Graph 1.**
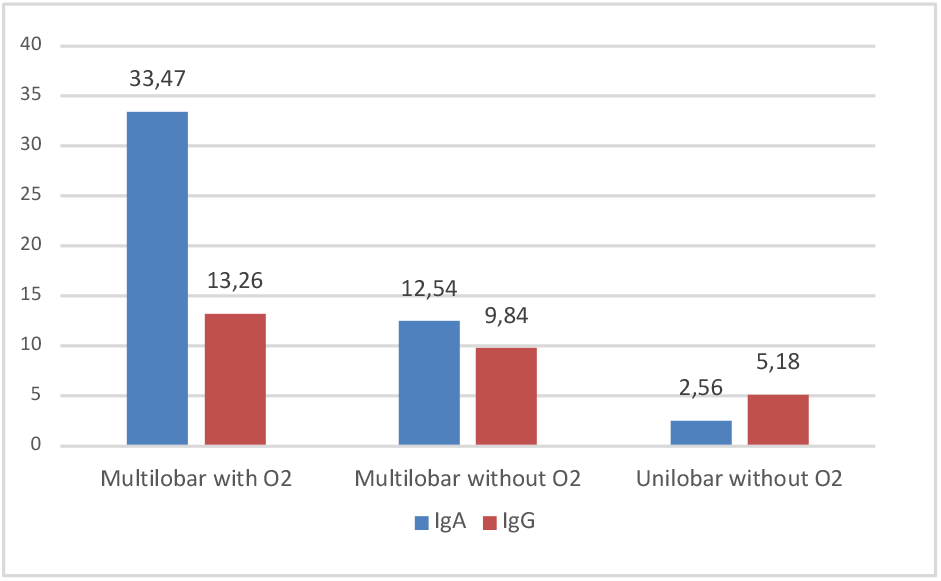
Antibody Titration x Severity

Previous studies with SARS showed that 52.4% of the patients who were hospitalized maintained abnormalities in the DLCO after 2 years (19).

However, in other analysis, the improvement of this parameter occurred in 80.4% in 3 months (20).

Seow J. et al showed a peak of IgM and IgA between 20 and 30 days, respectively, with a rapid drop after this interval. The levels of these antibodies are higher according to the severity of the case, which is not observed with IgG, remaining at similar levels regardless of severity. There was a decrease in the levels of neutralizing antibodies and of IgG in almost all patients within 3 months (21).

In our analysis, we had the same observation, with the most severe patients presenting higher levels of IgA and IgG. The gradual drop after the peak of antibody production was not the subject of our study.

Zhao et al analyzed patients who were hospitalized for viral pneumonia, confirmed by COVID 19 through RT-PCR, and submitted to specific serology at different intervals. The sensitivity of serological tests using the ELISA method is 38.3% in the first week, reaching 90% around the twelfth day. From the eighth to the fourteenth day, its sensitivity is 89.6%, 73.3%, and 54.1% for total antibodies, IgM, and IgG, respectively. When analyzing the results ≥ 15 days, the detection was 100%, 94.3%, and 79.8% for total antibodies, IgM, and IgG, respectively. The specificity was 100% (5). Our results confirm the study by Lou B. et al, in which 80 patients diagnosed with SARS-CoV-2 pneumonia were followed up, confirmed by RT-PCR associated with the presence of tomographic findings, who underwent specific serological tests. The sensitivity of total antibodies (Ab), IgM, and IgG from 0 to 7 days after the onset of symptoms was 64.1%, 33.3%, and 33.3%, respectively. When serology is analyzed after 2 weeks, sensitivity rises to Ab, 100%; IgM, 96.7%; and IgG, 93.3%. Seroconversion time was 9 days for Ab, 10 days for IgM, and 12 days for IgG, after symptom onset (14).

We also observed a proportional inversion in the IgA and IgG titers after the fourth week. A study evaluating the clinical and immunological characteristics showed that the levels of IgG and neutralizing antibodies decreased in 93.3% and 81.1% of asymptomatic patients, respectively, and 96.8% and 62.2% of symptomatic patients, respectively. The drop in the antibodies to normal levels occurred in 40% of asymptomatic patients and 12.9% of symptomatic patients within 2 to 3 months (22). It has also been characterized that symptomatic patients have higher levels of antibodies than asymptomatic ones. Another observation is that high levels of IgM in the acute phase of the disease correlate with inflammatory laboratory parameters, that is, the greater the inflammatory tests, the greater the level of IgM.

Kissler et al, using mathematical models and data extrapolated from infections by other types of coronavirus, suggested that if immunity against SARS-CoV-2 persists for approximately 40 weeks or 2 years, epidemic outbreaks would occur every 1 to 2 years, respectively. In the case of a permanent immune response, COVID 19 could be extinguished within an interval of up to 5 years (23). Such information reinforces the need to understand the immunological effect based on the acquired infection. Because they are people who are still working, the vast majority of individuals in our study belong to a lower age group - a median of 40.5 years.

According to Wang et al, the average age in hospitalizations for pneumonia is 56 years (22-92 years) (7).

Although men are predominant (9), our result showed a higher percentage of women due to the profile of employees at our institution.

We also found a difference in the immune response with IgA levels six times higher in men compared to women (P <0.05). Although it is known that there is the greatest risk of men evolving with severe forms and, therefore, higher antibody titers, our percentage of patients who collected IgA and required oxygen supplementation in men was 38% (3/8) and in women, 35% (6/17), respectively. Wu et al. also showed a greater immune response in males, characterized by high levels of neutralizing antibodies (24).

Symptoms such as dyspnea, cough, and fever are highly related to pneumonia. In our study, these symptoms were present in 88%, 85%, and 65% of patients, respectively. In the context of the pandemic, the combination of these symptoms often leads to the suspicion of COVID 19 with pulmonary impairment, requiring a chest CT scan. In our sample, most cases presented multilobar impairment at the time of diagnosis (85%), consistent with previous studies (25, 26).

Another topic of discussion is the time after the onset of symptoms that the RT-PCR would remain positive. The average time reported in the literature has been 20 days (8-37 days), not necessarily indicating the potential for contagion during this period (27). Studies based on viral culture have shown that the infectivity period starts 2 to 3 days before the clinical manifestation, extending up to a maximum of 8 days after the onset of symptoms in mild cases (28, 29). It is not possible to state that patients who have a severe form of the disease have the same transmission interval. According to our observation, 50% of patients who underwent a second RT-PCR collection had positive results, with a median collection of 31 days after symptom onset. According to our observation, patients who presented positive second RT-PCR with collection after 3 weeks did not have higher antibody titers compared to negative RT-PCR results, thus suggesting that positive persistent molecular analysis does not translate into greater immune response.

Transmission in a hospital environment has also been another subject of discussion. According to Wang et al., 29% of the cases that were confirmed with COVID 19 and viral pneumonia were healthcare professionals (7). In this study, 77.5% were from the inpatient unit, 17.5% from the emergency department, and 5% from the intensive care unit. Of the total number of employees evaluated in our sample, 65% worked in the Intensive Care Unit (ICU) - nursing, physiotherapists, and clerks. It is not possible to affirm that all individuals subject to the study had a focus on hospital contamination, but the high percentage of cases related to work in the ICU suggests that this environment is the most at risk for contagion in the care of patients with severe/critical COVID. A study carried out with Northwell Health System health professionals showed a prevalence of 13.7% in this population by using the serological test. Although 73% of the total are women, the prevalence was equivalent between genders, with a predominance of ICU employees and specific hospitalization units for COVID 19 (30). Asymptomatic and symptomatic patients seem to have a similar viral load, but with a longer clearance time in the latter group (31). Despite this, it is expected that patients admitted to the ICU are in the main phase of viral replication and, therefore, offer a higher risk of contagion. The frequent manipulation of patients, in addition to the greater elimination of droplets/aerosols, would be rational explanations for this result (32). We did not observe a significant difference in the percentage of individuals with multilobar pulmonary impairment and the levels of antibodies among ICU employees compared to employees in other sectors.

By the end of this study, we had approximately 1200 employees infected with the new coronavirus. Although several cases of physicians at the institution were diagnosed with COVID 19, few were the object of our study because they have a different flow of care. The main limitation of the study was the low sample of patients who presented the criteria for inclusion in this study. The difficulty in performing serological tests with the technique selected for evaluation ended up restricting the group of patients to be evaluated. Another limitation was the lack of quantification of viral load, making it impossible to establish a relationship between the antibody titer and the severity of the condition with the virus load in the body.

## 6 Conclusion

In patients who were diagnosed with COVID 19, hospitalized, with pulmonary manifestation confirmed by tomography, the serological test performed using the ELISA method showed 100% sensitivity and specificity as from 14 days after the onset of symptoms. Our results favor the performance of such an examination in patients who initially had repeatedly negative RT-PCR results, but with a pulmonary impairment which suggests SARS-CoV-2 infection. As pulmonary sequelae can be long-lasting, the etiological diagnosis of pneumopathy is of fundamental importance to understand the evolution of the disease and therapeutic suitability.

## Supporting information

ethics committee on research opinion

## Data Availability

All data is available to answer any questions

## References

1. Munster VJ, Koopmans M, van Doremalen N, van Riel D, de Wit E. A Novel Coronavirus Emerging in China - Key Questions for Impact Assessment. N Engl J Med. 2020;382(8):692–4. http://doi.org/10.1056/NEJMp2000929

2. WHO Coronavirus Disease (COVID-19) Dashboard. https://covid19.who.int. Date last updated: October 13 2020. Date last accessed: October 13 2020.

3. WHO. Clinical management of COVID 19. ovhttps://www.who.int/publications/i/item/clinical-management-of-covid-19. Date last updated: May 27 2020. Date last accessed: October 13 2020.

4. Xie J, Ding C, Li J, Wang Y, Guo H, Lu Z, et al. Characteristics of patients with coronavirus disease (COVID-19) confirmed using an IgM-IgG antibody test. J Med Virol. 2020. http://doi.org/10.1002/jmv.25930

5. Zhao J, Yuan Q, Wang H, Liu W, Liao X, Su Y, et al. Antibody responses to SARS-CoV-2 in patients of novel coronavirus disease 2019. Clin Infect Dis. 2020. http://doi.org/10.1093/cid/ciaa344

6. Ai J-W, Zhang H-C, Xu T, Wu J, Zhu M, Yu Y-Q, et al. Optimizing diagnostic strategy for novel coronavirus pneumonia, a multi-center study in Eastern China. medRxiv. 2020:2020.02.13.20022673. https://doi.org/10.1101/2020.02.13.20022673

7. Wang D, Hu B, Hu C, Zhu F, Liu X, Zhang J, et al. Clinical Characteristics of 138 Hospitalized Patients With 2019 Novel Coronavirus-Infected Pneumonia in Wuhan, China. JAMA. 2020. http://doi.org/10.1001/jama.2020.1585

8. Chate RC, Fonseca E, Passos RBD, Teles G, Shoji H, Szarf G. Presentation of pulmonary infection on CT in COVID-19: initial experience in Brazil. J Bras Pneumol. 2020;46(2):e20200121.

9. Lai CC, Shih TP, Ko WC, Tang HJ, Hsueh PR. Severe acute respiratory syndrome coronavirus 2 (SARS-CoV-2) and coronavirus disease-2019 (COVID-19): The epidemic and the challenges. Int J Antimicrob Agents. 2020;55(3):105924. http://doi.org/10.1016/j.ijantimicag.2020.105924

10. Zhang JJ, Dong X, Cao YY, Yuan YD, Yang YB, Yan YQ, et al. Clinical characteristics of 140 patients infected with SARS-CoV-2 in Wuhan, China. Allergy. 2020. https://doi.org/10.1111/all.14238

11. Danrad R, Smith DL, Kerut EK. Radiologic chest CT findings from COVID-19 in Orleans Parish, Louisiana. Echocardiography. 2020;37(4):628–31. https://doi.org/10.1111/echo.14662

12. Lisboa Bastos M, Tavaziva G, Abidi SK, Campbell JR, Haraoui LP, Johnston JC, et al. Diagnostic accuracy of serological tests for covid-19: systematic review and meta-analysis. BMJ. 2020;370:m2516. https://doi.org/10.1136/bmj.m2516

13. Deeks JJ, Dinnes J, Takwoingi Y, Davenport C, Spijker R, Taylor-Phillips S, et al. Antibody tests for identification of current and past infection with SARS-CoV-2. Cochrane Database Syst Rev. 2020;6:CD013652. https://doi.org/10.1002/14651858.CD013652

14. Lou B, Li TD, Zheng SF, Su YY, Li ZY, Liu W, et al. Serology characteristics of SARS-CoV-2 infection since exposure and post symptom onset. Eur Respir J. 2020. http://doi.org/10.1183/13993003.00763-2020

15. Ai T, Yang Z, Hou H, Zhan C, Chen C, Lv W, et al. Correlation of Chest CT and RT-PCR Testing in Coronavirus Disease 2019 (COVID-19) in China: A Report of 1014 Cases. Radiology. 2020:200642. https://doi.org/10.1148/radiol.2020200642

16. Yang Y, Yang M, Shen C, Wang F, Yuan J, Li J, et al. Evaluating the accuracy of different respiratory specimens in the laboratory diagnosis and monitoring the viral shedding of 2019-nCoV infections. medRxiv. 2020:2020.02.11.20021493. https://doi.org/10.1101/2020.02.11.20021493

17. Imai K, Tabata S, Ikeda M, Noguchi S, Kitagawa Y, Matuoka M, et al. Clinical evaluation of an immunochromatographic IgM/IgG antibody assay and chest computed tomography for the diagnosis of COVID-19. J Clin Virol. 2020;128:104393. https://doi.org/10.1016/j.jcv.2020.104393

18. Mo X, Jian W, Su Z, Chen M, Peng H, Peng P, et al. Abnormal pulmonary function in COVID-19 patients at time of hospital discharge. Eur Respir J. 2020;55(6). https://doi.org/10.1183/13993003.01217-2020

19. Ngai JC, Ko FW, Ng SS, To KW, Tong M, Hui DS. The long-term impact of severe acute respiratory syndrome on pulmonary function, exercise capacity and health status. Respirology. 2010;15(3):543–50. https://doi.org/10.1111/j.1440-1843.2010.01720.x

20. Xie L, Liu Y, Xiao Y, Tian Q, Fan B, Zhao H, et al. Follow-up study on pulmonary function and lung radiographic changes in rehabilitating severe acute respiratory syndrome patients after discharge. Chest. 2005;127(6):2119–24. https://doi.org/10.1378/chest.127.6.2119

21. Seow J, Graham C, Merrick B, Acors S, Steel KJA, Hemmings O, et al. Longitudinal evaluation and decline of antibody responses in SARS-CoV-2 infection. medRxiv. 2020:2020.07.09.20148429. https://doi.org/10.1101/2020.07.09.20148429

22. Long QX, Tang XJ, Shi QL, Li Q, Deng HJ, Yuan J, et al. Clinical and immunological assessment of asymptomatic SARS-CoV-2 infections. Nat Med. 2020. https://doi.org/10.1038/s41591-020-0965-6

23. Kissler SM, Tedijanto C, Goldstein E, Grad YH, Lipsitch M. Projecting the transmission dynamics of SARS-CoV-2 through the postpandemic period. Science. 2020;368(6493):860–8. http://doi.org/10.1126/science.abb5793

24. Wu F, Liu M, Wang A, Lu L, Wang Q, Gu C, et al. Evaluating the Association of Clinical Characteristics With Neutralizing Antibody Levels in Patients Who Have Recovered From Mild COVID-19 in Shanghai, China. JAMA Internal Medicine. 2020. https://doi.org/10.1001/jamainternmed.2020.4616

25. Zhu J, Zhong Z, Li H, Ji P, Pang J, Li B, et al. CT imaging features of 4121 patients with COVID-19: A meta-analysis. J Med Virol. 2020;92(7):891–902. https://doi.org/10.1002/jmv.25910

26. Sun Z, Zhang N, Li Y, Xu X. A systematic review of chest imaging findings in COVID-Quant Imaging Med Surg. 2020;10(5):1058–79. http://doi.org/10.21037/qims-20-564

27. Zhou F, Yu T, Du R, Fan G, Liu Y, Liu Z, et al. Clinical course and risk factors for mortality of adult inpatients with COVID-19 in Wuhan, China: a retrospective cohort study. Lancet. 2020;395(10229):1054–62. https://doi.org/10.1016/S0140-6736(20)30566-3

28. He X, Lau EHY, Wu P, Deng X, Wang J, Hao X, et al. Temporal dynamics in viral shedding and transmissibility of COVID-19. Nat Med. 2020;26(5):672–5. https://doi.org/10.1038/s41591-020-0869-5

29. Wolfel R, Corman VM, Guggemos W, Seilmaier M, Zange S, Muller MA, et al. Virological assessment of hospitalized patients with COVID-2019. Nature. 2020;581(7809):465–9. https://doi.org/10.1038/s41586-020-2196-x

30. Moscola J, Sembajwe G, Jarrett M, Farber B, Chang T, McGinn T, et al. Prevalence of SARS-CoV-2 Antibodies in Health Care Personnel in the New York City Area. JAMA. 2020. http://doi.org/10.1001/jama.2020.14765

31. Lee S, Kim T, Lee E, Lee C, Kim H, Rhee H, et al. Clinical Course and Molecular Viral Shedding Among Asymptomatic and Symptomatic Patients With SARS-CoV-2 Infection in a Community Treatment Center in the Republic of Korea. JAMA Internal Medicine. 2020. http://doi.org/10.1001/jamainternmed.2020.3862

32. Goh KJ, Wong J, Tien JC, Ng SY, Duu Wen S, Phua GC, et al. Preparing your intensive care unit for the COVID-19 pandemic: practical considerations and strategies. Crit Care. 2020;24(1):215. https://doi.org/10.1186/s13054-020-02916-4

