## Supplementary material for "Importance of serological testing in the convalescence phase in patients with pulmonary impairment due to COVID 19 - a health care workers analysis": ethics committee on research opinion

AMENDMENT DATA
Research Title: Importance of serological testing in the convalescence phase in patients with pulmonary involvement by COVID 19 - new gold standard?

Researcher: José Rodrigues Pereira
Thematic Area:
Version: 2
CAAE: 36217520.0.0000.5483
Proponent Institution: Real e Benemérita Associação Portuguesa de Beneficência / SP
Main Sponsor: Own Financing
OPINION DATA
Opinion Number: 4,240,541

Project presentation:

Summary:
After the discovery of a new coronavirus (SARS-CoV-2) in December 2019, more than 16 million cases were confirmed worldwide and more than 650 thousand deaths were related to complications from this disease (COVID 19). Since then, all the nuances related to the virus, such as understanding the forms of contagion, the characteristics of the symptoms, the degree of impairment, the diagnostic and therapeutic approach have been
studied. Based on clinical suspicion, the diagnostic method of choice is molecular analysis through RT PCR, using material from nasopharyngeal and oropharyngeal swab. According to information from scientific literature, its sensitivity is around 70%.
During the course of the disease, the lung is the most compromised organ and depending on the severity, the sequelae can be long-lasting and permanent.
Due to the possibility of lasting pulmonary impairment and the limited sensitivity of RT PCR, there will be cases where the patient will be submitted to medical evaluation with respiratory symptoms and pulmonary impairment of unknown etiology.
Serological tests have low sensitivity, approximately 30%, for the diagnosis of COVID in the 1st week after the onset of symptoms, however they may have an epidemiological relevance in patients with pulmonary impairment of unknown etiology.
In those cases, they may be an
useful tool in the etiological diagnosis of pulmonary diseases with interstitial involvement.
Another current question is whether the persistence of specific antibodies would result in lasting immunity against SARS-CoV-2. Studies have shown gradual decline over the months, with the possibility of normalization and risk of reinfection in case of new exposure. Such doubt also extends to the vaccines that are being tested to date.
Understanding the immunological curve as well as the profile of patients who have a greater response is necessary to adapt research and safety guidelines.
Our study will evaluate the sensitivity and specificity of serological methods during the convalescence phase of patients hospitalized with suspected or confirmed COVID 19 and viral pneumonia.

All patients evaluated are health professionals, employees of the Beneficência Portuguesa Hospital of São Paulo. Our secondary analysis will evaluate the value of serological titers according to the severity of symptoms, workplace, sex and comorbidities, in order to identify a profile of patients who would have a greater immune response and probably a longer time of active immunization against SARS-CoV-2.

Research Objective:

Primary Outcome:
Analysis of sensitivity and specificity of the serological test as a diagnostic tool in patients with COVID 19 with pulmonary impairment in the convalescence phase.

Secondary outcome:
Analyze the values of the serological titration according to the severity of the pulmonary involvement by SARS-CoV-2, sex and variability according to the collection time in health professionals.

Assessment of Risks and Benefits:
Risks:
There are no risks involved
Benefits:
Demonstrate the importance of the serological test for COVID 19 in the convalescence phase, with the suggestion of a new gold standard in diagnosis in patients with negative RT PCR. Understand immunological behavior, through the titers of IgA and IgG, according to sex and disease severity.

Research Comments and Considerations:

This researcher's cover page and brochure were presented in this amendment.
This is a retrospective study, evaluating the effectiveness of serological tests in employees of BP´Beneficência Portuguesa de São Paulo who had COVID-19 confirmed by RT-PCR.

Mandatory submission terms considerations:

Exemption from the informed consent was requested, due to the retrospective nature of the study.

Conclusions or Pending and List of Inadequacies:
Approved without pending issues.

Final Considerations at the discretion of the CEP:
In view of the above, the Ethics Committee in Research of the Portuguese Beneficence, according to the attributions defined in Resolution CNS No. 466/2012 and subsequent ones, manifests itself by the Project Approval, as proposed for the beginning of the Research.
We request that semiannual reports on the progress of the research should be presented to this CEP, as well as information related to the modifications of the protocol, cancellation, closure and destination of the knowledge obtained.
